# Open science policies of medical and health sciences journals before and during the COVID-19 pandemic: a repeat cross-sectional study

**DOI:** 10.1101/2022.01.26.22269868

**Authors:** Antoni D. Gardener, Ellen J. Hicks, Chloe Jacklin, Gifford Tan, Aidan G. Cashin, Hopin Lee, David Nunan, Elaine C. Toomey, Georgia C. Richards

## Abstract

Cross-disciplinary openness and transparency of research plays an important role in scientific progress. We evaluated open-science related policies of 19 high ranking health and medical journals before (February 2020) and during (May 2021) the COVID-19 pandemic. The Transparency and Openness Promotion (TOP) guideline and the International Committee of Medical Journal Editors (ICMJE) requirements for disclosing conflicts of interest (COIs) were used to audit journal policies. TOP scores slightly improved during the COVID-19 pandemic, from a median of 5 (IQR: 2-12.5) out of a possible 24 points in February 2020 to 7 (IQR: 4-12) in May 2021. Most journals fulfilled all ICMJE provisions for reporting COIs before (84%; n=16) and during (95%; n=18) the COVID-19 pandemic. The COVID-19 pandemic has highlighted the importance of practising open science, however adherence to open science standards in audited policies was low overall, which may reduce progress in health and medical research.

## INTRODUCTION

Scientific advancement in any field or discipline relies considerably on transparent research practices^1^. This includes medicine and public health. The ability to access, evaluate, and extrapolate the findings from research to clinical practice and policy relies on the transparency of available evidence^1,2^. Research transparency is also imperative for the replication of scientific findings^1^. Given the often high-stakes circumstances of evidence-based medical and public health practice, the need for transparency is particularly critical.

Journal policies that support open science practices, also called open research or open scholarship, have improved transparency and replicability^3,4^. Open science is defined as research activities and outputs that are made publically available with little to no restrictions^5^. Open and transparent practices include, but are not limited to, the preregistration of study protocols and analysis plans, the sharing of data, statistical code, and research materials, and the disclosure of authors’ conflicts of interests. Making code and data openly available allows for reuse, replication, and peer assessment, which is an important step in verifying the findings of studies and progressing science^6^. Poor adherence to open practices produces unreliable outcomes and slows scientific progress^7–9^.

Research published during the COVID-19 pandemic has highlighted the widespread impact of practising ‘closed’ research, where cases of undisclosed data and a lack of transparency led to high-profile retractions^10,11^. This can compromise public trust, sway public opinions and policies, and bias future research^12^. The pandemic has also demonstrated the positive impacts of open sciences practices, as demonstrated by quick data sharing used to identify the COVID-19 genome rapidly and journals removing paywalls for COVID-19 research, resulting in more equitable and widespread access to research^13–15^.

There are various tools available to support and reward the practice of open science, including freely available repositories (e.g. GitHub, Figshare) and platforms (e.g. the Open Science Framework [OSF]), publication methods (e.g. registered reports), and open science badges^16–19^. However, some researchers have expressed concerns regarding practising openly, including but not limited to one’s intellectual property not being cited or acknowledged correctly, losing a competitive advantage to groups working on similar research, practicality concerns, and patient confidentiality^20,21^. Other factors that are likely to contribute to poor adherence include a lack of knowledge or skills in using these tools to facilitate openness and the flawed incentive system that encourages the publication of more instead of better research^22,23^. For transparent research practices to be the default, stakeholders, including institutions, funders, publishers, and journals, can mandate and facilitate researchers to work in such ways. Since journals are the gatekeepers of disseminating research, strengthening journal policies to reflect transparent and open practices could incentivise researchers to upskill and practice open science.

While efforts have been made to improve and promote the transparency of research^1,22^, further steps are needed to adopt and normalise this way of working. Preregistration of studies, analysis plans, and registered reports are valuable ways of limiting publication bias and misleading practices such as HARKing (Hypothesising After the Results are Known) or p-hacking^24–26^. The Centre for Open Science (COS), which has pioneered the development of open tools and resources, developed the Transparency and Openness Promotion (TOP) guideline in 2015 to assess and improve journal policy requirements of publishing open and transparent research^27^. Similar policy changes have improved open and transparent research practices, as was seen after the ICMJE trial registration policy in 2004^28^. Since the publication of the TOP guideline, several evaluations of journal policies using the TOP guidelines have been published^29–31^, which suggest poor compliance to TOP guideline standards across multiple fields of research.

Since the beginning of the COVID-19 pandemic, several journals have released statements and made efforts to improve the openness and transparency of COVID-19 research to support researchers during the pandemic^32,33^. To date, a peer-reviewed evaluation of high-ranking general medical and health science journals assessing open science practices before or during the COVID-19 pandemic has not been conducted. Therefore, we aimed to conduct this evaluation and explore the potential impact of the COVID-19 pandemic on the adherence to transparent and open science standards of leading health and medical journals.

## ONLINE METHODS

We designed a cross-sectional study based on methods developed by Cashin et al.^29^ and preregistered the study protocol on the OSF^34^. We completed the first policy audit in early 2020 and repeated the same audit in 2021 to assess the potential impact of the COVID-19 pandemic on journal policies.

### Sampling procedure

On 31 January 2020, we sampled 19 journals listed in Google Scholar’s Top Publications for the health and medical sciences subcategory^35^, which at this time were (in alphabetical order): *Blood, Cell, Circulation, European Heart Journal, Gastroenterology, Journal of Clinical Oncology, Journal of the American College of Cardiology, Nature Genetics, Nature Medicine, Nature Neuroscience, Neuron, PLoS ONE, Proceedings of the National Academy of Sciences (PNAS), Science Translational Medicine, The British Medical Journal (BMJ), The Journal of the American Medical Association (JAMA), The Lancet, The Lancet Oncology*, and *The New England Journal of Medicine (NEJM)*. On 10 February 2020, the policies for each journal were downloaded and saved with date and time stamps, defined as the pre-COVID-19 pandemic timepoint. We repeated this procedure for each journal to collect updated policies between 26 and 31 May 2021, defined as during the COVID-19 pandemic timepoint. All policies extracted are openly available on our OSF repository^36^.

### Measurement tools and scoring

We used the TOP guideline to audit and score each journal’s policy. All audits were performed in duplicate by two independent study authors (ADG, EH, CJ, GT, AGC, HL, DN, ET), and discrepancies were adjudicated with a third author (GCR). The TOP guideline has eight standards: data citation, data transparency, code transparency, materials transparency, design and analysis transparency, study preregistration, analysis preregistration, and replication^27,37^. For scoring, we used version 1 of the TOP rubric^38^, where 0 (zero) denotes “no mention” (of the standard in the policy wording), 1 “discloses”, 2 “requires”, and 3 “verifies”. The maximum score obtainable for TOP was 24. Justifications for scoring, along with quoted material from the journal policy, where appropriate, are provided in the Google Sheet master files on our OSF repository^36^. Where a policy demonstrated varying requirements (for example, different types of study design) or only mentioned requirements for one element (for example, requiring the use of reporting guidelines or study preregistration for clinical trials only), a score was given based on the lowest-scoring element in accordance with the TOP guidelines.

Journals and funders can choose to become signatories of TOP to express their support, interest, and commitment in adopting such standards^37^. Hence we also scored the included journals on whether they were a signatory of TOP, permitted registered reports, and used open science badges. For these metrics, we provided scores based on previously published methods (i.e. TOP signatory: no=0, yes=1; registered reports: no=0, yes=2; and open science badges: no=0, yes=2)^29^.

To examine the journal’s requirements for authors to disclose conflict of interests (COIs), we used the four standards in the International Committee of Medical Journal Editors (ICMJE) disclosure form^39^. The four standards include: 1) recipient of payment or services from a third party (government, commercial, private foundation) for any aspect of the submitted work; 2) any financial relationships (regardless of the amount of compensation) with entities in the biomedical arena that could be perceived to influence, or that give the appearance of influencing, the submitted work; 3) any patents, whether planned, pending or issued, broadly relevant to the submitted work; and 4) any other relationships or activities that readers could perceive to have influenced, or that give the appearance of potentially influencing, the submitted work. We scored the journal policies for disclosing COIs on a 0 to 4 scale, with 0 being no statement of any of the standards and four being all standards or provision of the complete ICMJE disclosure form in accordance with published methods.^24^

## Data analysis

All scores were provided in separate Google Sheets that were combined, compared, and converted to a .csv file for analysis. We used descriptive statistics and calculated medians and interquartile ranges (IQRs) of scores where appropriate. Journals were ranked by their scores for the eight TOP standards (out of 24) and requirements for COIs (out of 4). To assess where journals engaged most with TOP standards, we summed scores across journals for each TOP standard, where the maximum total could be 57 (3 possible points for 19 journals). Scores were compared before and during the COVID-19 pandemic using absolute values and percentage changes.

### Software and data sharing

We used pandas, seaborn, and matplotlib modules in Jupyter Notebooks with Python v3.7 to analyze the scores and create figures. Our study materials, data, code, and figures are openly accessible on our OSF repository^36^ and GitHub^40^.

### Protocol deviations

In the protocol, we planned to examine the 20 journals listed in Google Scholar’s Top Publications for health and medical sciences as listed on the search date (31 January 2020). We excluded the *Cochrane Database of Systematic Reviews* during analysis as it is a specialized journal that commissions research groups to adhere to a prescriptive methodology and differs significantly from the other included journals. Therefore, our study includes 19 journals. We preregistered a follow-up study to assess the impact of informing journals of their TOP scores in March 2020^41^. However, this second study was not conducted owing to the onset of the COVID-19 pandemic and the priority for journals to review and disseminate pressing COVID-19 related research. Instead, we repeated our evaluation in May 2021 to assess the adherence to the TOP guidelines during the COVID-19 pandemic, but did not pre-register the plans for this follow-up evaluation.

## RESULTS

For the eight TOP standards, journals had a median score of 5 (IQR: 2-12.5) out of a possible maximum score of 24 before the COVID-19 pandemic (February 2020), which increased to a median of 7 (IQR: 4-12) in May 2021. The highest scoring journals were *Nature Medicine* and *Nature Neuroscience*, with TOP scores of 14/24 for both evaluations (Figure 1). The *Journal of the American College of Cardiology* had the lowest score at each time point, receiving a TOP score of 0/24.

**Figure 1:**
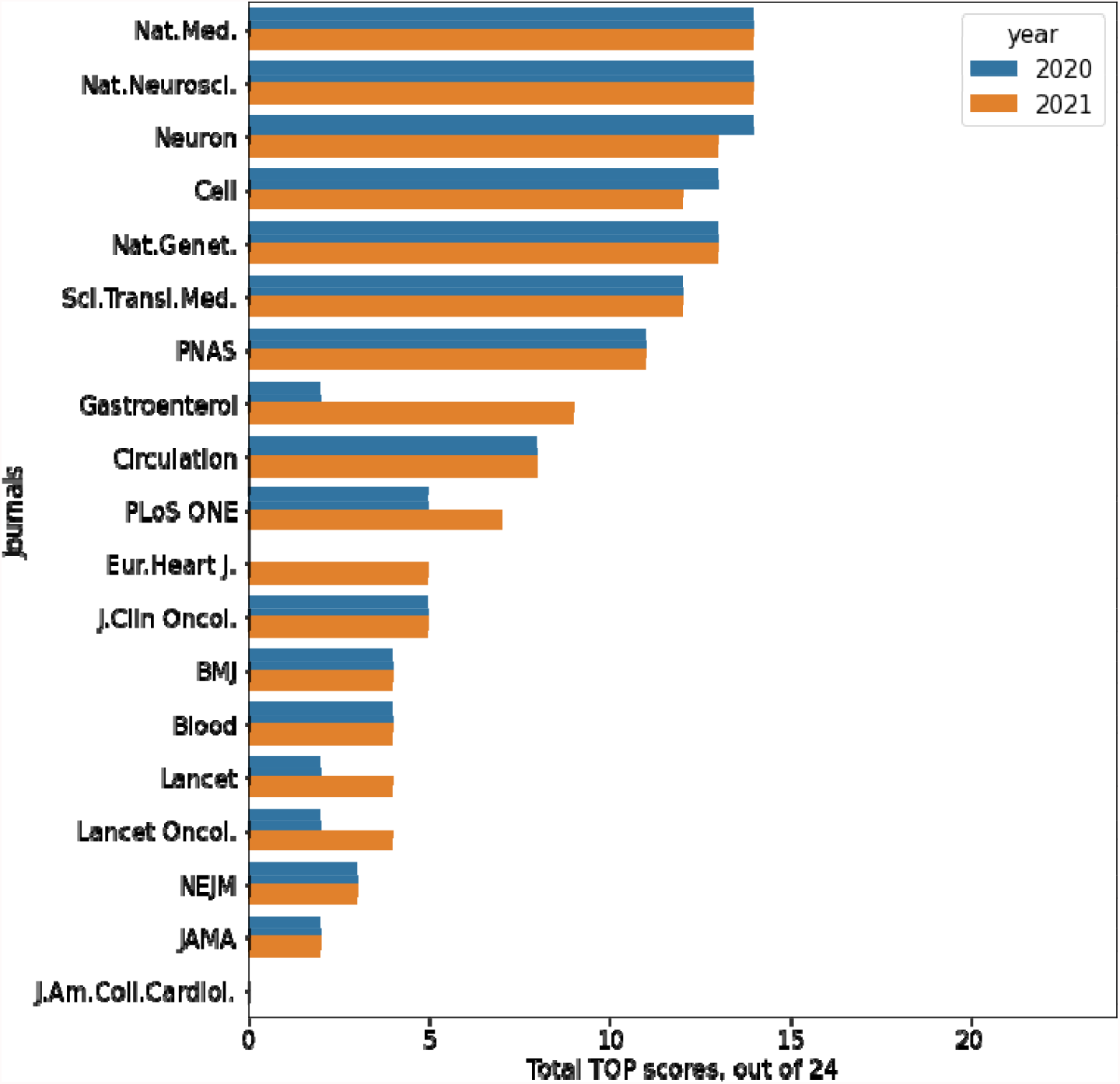
Scores for 19 high-ranking health and medical sciences journal policies using the Transparency and Openness Promotion (TOP) guidelines in descending order for 2021 scores. The first audit was conducted on policies dated in February 2020 and the second set of policies were from May 2021. Policies had a median score of 5 (IQR: 2-12.5) out of a possible maximum score of 24 in February 2020, which increased to a median of 7 (IQR: 4-12) in May 2021. Eur.Heart J: European Heart Journal; Gastroenterol: Gastroenterology; J.Clin Oncol: Journal of Clinical Oncology; J.Am.Coll.Cardiol: Journal of the American College of Cardiology; Nat.Genet: Nature Genetics, Nat.Med: Nature Medicine; Nat.Neurosci: Nature Neuroscience; PNAS: Proceedings of the National Academy of Sciences; Sci.Transl.Med: Science Translational Medicine; BMJ: The British Medical Journal; JAMA: Journal of the American Medical Association; Lancet Oncol: The Lancet Oncology; and NEJM: The New England Journal of Medicine.

Eleven journals (58%) had no change in their TOP score following the onset of the COVID-19 pandemic. One-quarter of journals (26%, n=5) improved their policies; *Gasterenrology* had the greatest improvement (from a score of 2 to a score of 9), followed by the *European Heart Journal* (score of 0 to a score of 5), with the remaining three journals improving by a score of 2 (*The Lancet, The Lancet Oncology*, and *PLoS ONE*). Two journals, *Cell* and *Neuron*, had a reduction in their TOP score during the COVID-19 pandemic (Figure 1).

Across the 19 medical and health sciences journal policies, the sum of scores for each TOP standard improved on average during the COVID-19 pandemic (May 2021) compared with before (Februrary 2020; Figure 2). Journal policies scored highest for their adherence to data transparency and design and analysis transparency. Policies engaged less with the requirements for preregistration of study protocols and analysis plans and the submission of replication studies. The scores for each TOP standard are summarised in Supplement 1 and Table 1.

**Figure 2:**
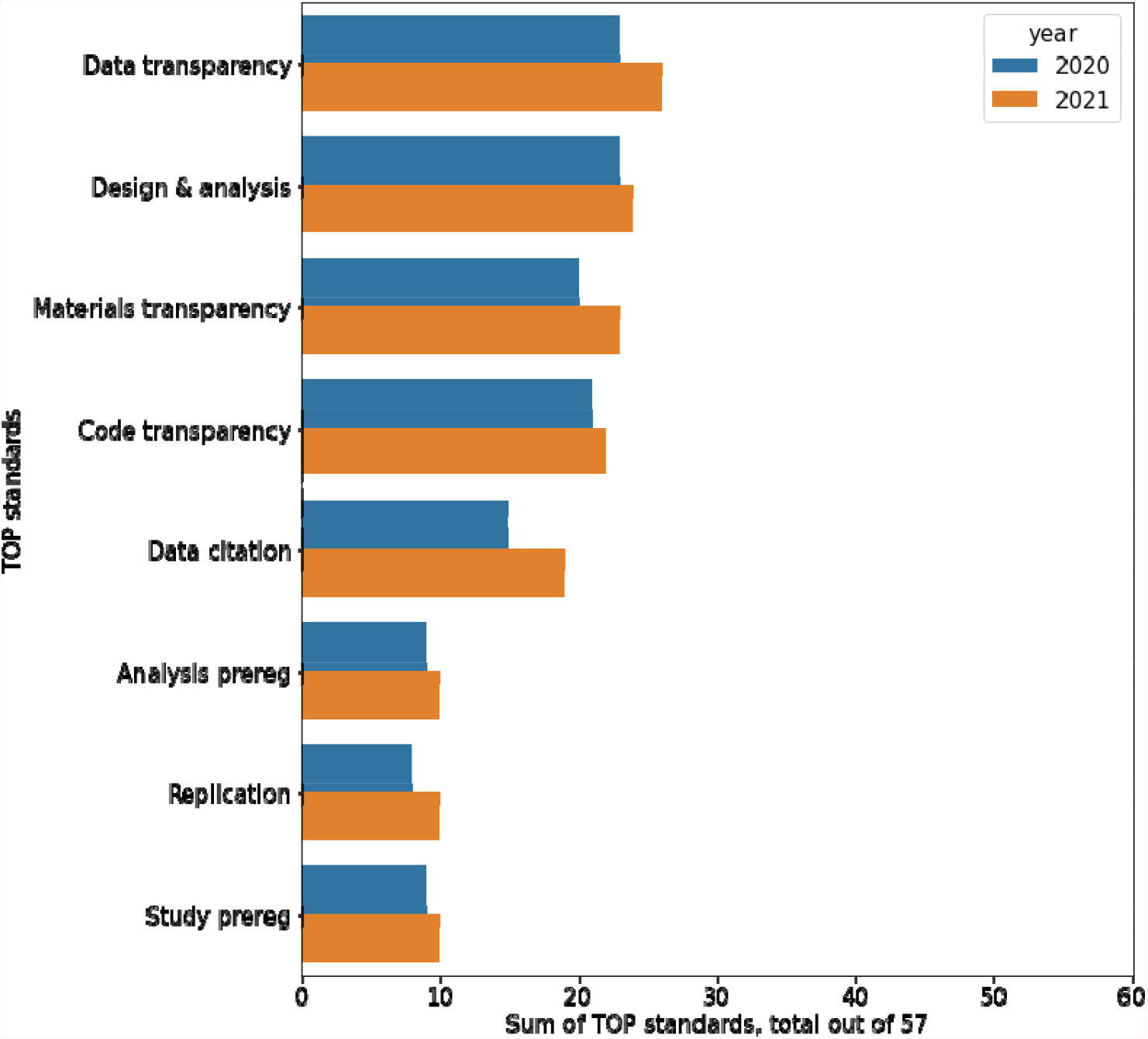
Cumulative scores on the eight Transparency and Openness Promotion (TOP) standards for 19 medical and health sciences journals evaluated before the COVID-19 pandemic (February 2020) and during the COVID-19 pandemic (May 2021) in descending order using 2021 scores.

**Table 1:** Scores for the journal policies of 19 health and medical sciences journals using the Transparency and Openness Promotion (TOP) guideline before the COVID-19 pandemic (February 2020) and during (May 2021), ordered alphabetically by Journal.

| Journal | Transparency |  |  |  |  |  |  |  |  |  |  |  |  |  |  |  |  |  |  |  |
| --- | --- | --- | --- | --- | --- | --- | --- | --- | --- | --- | --- | --- | --- | --- | --- | --- | --- | --- | --- | --- |
|  | Citation |  | Data |  | Code |  | Materials |  | Design & analysis |  | Study prereg |  | Analysis prereg |  | Replication |  | TOP signatory |  | Reg reports | OS badges |
|  | 2020 | 2021 | 2020 | 2021 | 2020 | 2021 | 2020 | 2021 | 2020 | 2021 | 2020 | 2021 | 2020 | 2021 | 2020 | 2021 | 2020 | 2021 | 2020-21* | 2020-21* |
| Blood | 0 | 0 | 2 | 2 | 0 | 0 | 2 | 2 | 0 | 0 | 0 | 0 | 0 | 0 | 0 | 0 | 0 | 0 | 0 | 0 |
| BMJ | 0 | 0 | 1 | 1 | 0 | 0 | 0 | 0 | 2 | 2 | 1 | 1 | 0 | 0 | 0 | 0 | 1 | 1 | 0 | 0 |
| Cell | 2 | 2 | 2 | 2 | 3 | 2 | 1 | 1 | 2 | 2 | 1 | 1 | 1 | 1 | 1 | 1 | 1 | 1 | 0 | 0 |
| Circulation | 1 | 1 | 1 | 1 | 1 | 1 | 1 | 1 | 1 | 1 | 1 | 1 | 1 | 1 | 1 | 1 | 1 | 1 | 0 | 0 |
| European Heart Journal | 0 | 2 | 0 | 1 | 0 | 1 | 0 | 1 | 0 | 0 | 0 | 0 | 0 | 0 | 0 | 0 | 0 | 0 | 0 | 0 |
| Gastroente | 0 | 2 | 1 | 2 | 0 | 1 | 1 | 1 | 0 | 1 | 0 | 1 | 0 | 1 | 0 | 1 | 0 | 0 | 0 | 0 |
| J AM Coll Cardiol | 0 | 0 | 0 | 0 | 0 | 0 | 0 | 0 | 0 | 0 | 0 | 0 | 0 | 0 | 0 | 0 | 0 | 0 | 0 | 0 |
| J Clin Oncol | 0 | 0 | 1 | 1 | 2 | 2 | 2 | 2 | 0 | 0 | 0 | 0 | 0 | 0 | 0 | 0 | 0 | 0 | 0 | 0 |
| JAMA | 0 | 0 | 0 | 0 | 0 | 0 | 0 | 0 | 2 | 2 | 0 | 0 | 0 | 0 | 0 | 0 | 0 | 0 | 0 | 0 |
| Lancet | 0 | 0 | 0 | 1 | 0 | 0 | 0 | 1 | 2 | 2 | 0 | 0 | 0 | 0 | 0 | 0 | 0 | 0 | 0 | 0 |
| Lancet Oncol. | 0 | 0 | 0 | 1 | 0 | 0 | 0 | 1 | 2 | 2 | 0 | 0 | 0 | 0 | 0 | 0 | 0 | 0 | 0 | 0 |
| Nature Genetics | 2 | 2 | 2 | 2 | 2 | 2 | 2 | 2 | 1 | 2 | 1 | 1 | 1 | 1 | 1 | 1 | 1 | 1 | 0 | 0 |
| Nature Medicine | 2 | 2 | 2 | 2 | 3 | 3 | 2 | 2 | 2 | 2 | 1 | 1 | 1 | 1 | 1 | 1 | 1 | 1 | 0 | 0 |
| Nature Neuroscience | 2 | 2 | 2 | 2 | 3 | 3 | 2 | 2 | 2 | 2 | 1 | 1 | 1 | 1 | 1 | 1 | 1 | 1 | 0 | 0 |
| NEJM | 0 | 0 | 1 | 1 | 0 | 0 | 0 | 0 | 1 | 1 | 0 | 0 | 1 | 1 | 0 | 0 | 0 | 0 | 0 | 0 |
| Neuron | 2 | 2 | 2 | 2 | 3 | 2 | 2 | 2 | 2 | 2 | 1 | 1 | 1 | 1 | 1 | 1 | 1 | 1 | 0 | 0 |
| PLoS ONE | 1 | 1 | 2 | 2 | 0 | 1 | 1 | 1 | 1 | 1 | 0 | 0 | 0 | 0 | 0 | 1 | 1 | 1 | 2 | 0 |
| PNAS | 1 | 1 | 2 | 2 | 2 | 2 | 2 | 2 | 1 | 1 | 1 | 1 | 1 | 1 | 1 | 1 | 0 | 1 | 0 | 0 |
| Sci Transl Medicine | 2 | 2 | 2 | 2 | 2 | 2 | 2 | 2 | 1 | 1 | 1 | 1 | 1 | 1 | 1 | 1 | 1 | 1 | 0 | 0 |

\*identical scores were found in policies from February 2020 and May 2021. BMJ: The British Medical Journal; Gastroenterol: Gastroenterology; J Clin Oncol: Journal of Clinical Oncology; J Am Coll Cardiol: Journal of the American College of Cardiology; JAMA: Journal of the American Medical Association; Lancet Oncol: The Lancet Oncology; OS: open science; NEJM: The New England Journal of Medicine; PNAS: Proceedings of the National Academy of Sciences; Prereg: preregistration; Sci.Transl.Med: Science Translational Medicine; TOP: Transparency and Openness Promotion.

### Additional measures

Before the COVID-19 pandemic, nine journals were signatories of TOP, which increased to 10 journals in May 2021 after *PNAS* joined as a TOP signatory. *PLoS One* was the only journal that accepted registered reports at both time points. No journals used open science badges to acknowledge the authors’ efforts to preregister their study or openly share their data, code, and study materials.

## Data Availability

All data produced are available online at

https://osf.io/h2xud/

https://github.com/georgiarichards/TOP_medjournals

## Conflicts of interests

The median ICMJE score of reporting COIs was 4 (IQR: 4-4) at both time points. Before the COVID-19 pandemic (February 2020), 84% (16/19) of journals fulfilled all four ICMJE standards for reporting COIs (Figure 3). In May 2021, this increased to 95% (18/19). *Cell* was the only journal that did not fulfil the fourth ICMJE criteria across both time periods, which requires authors to report “any other relationships or activities that readers could perceive to have influenced, or that give the appearance of potentially influencing, the submitted work.”

**Figure 3:**
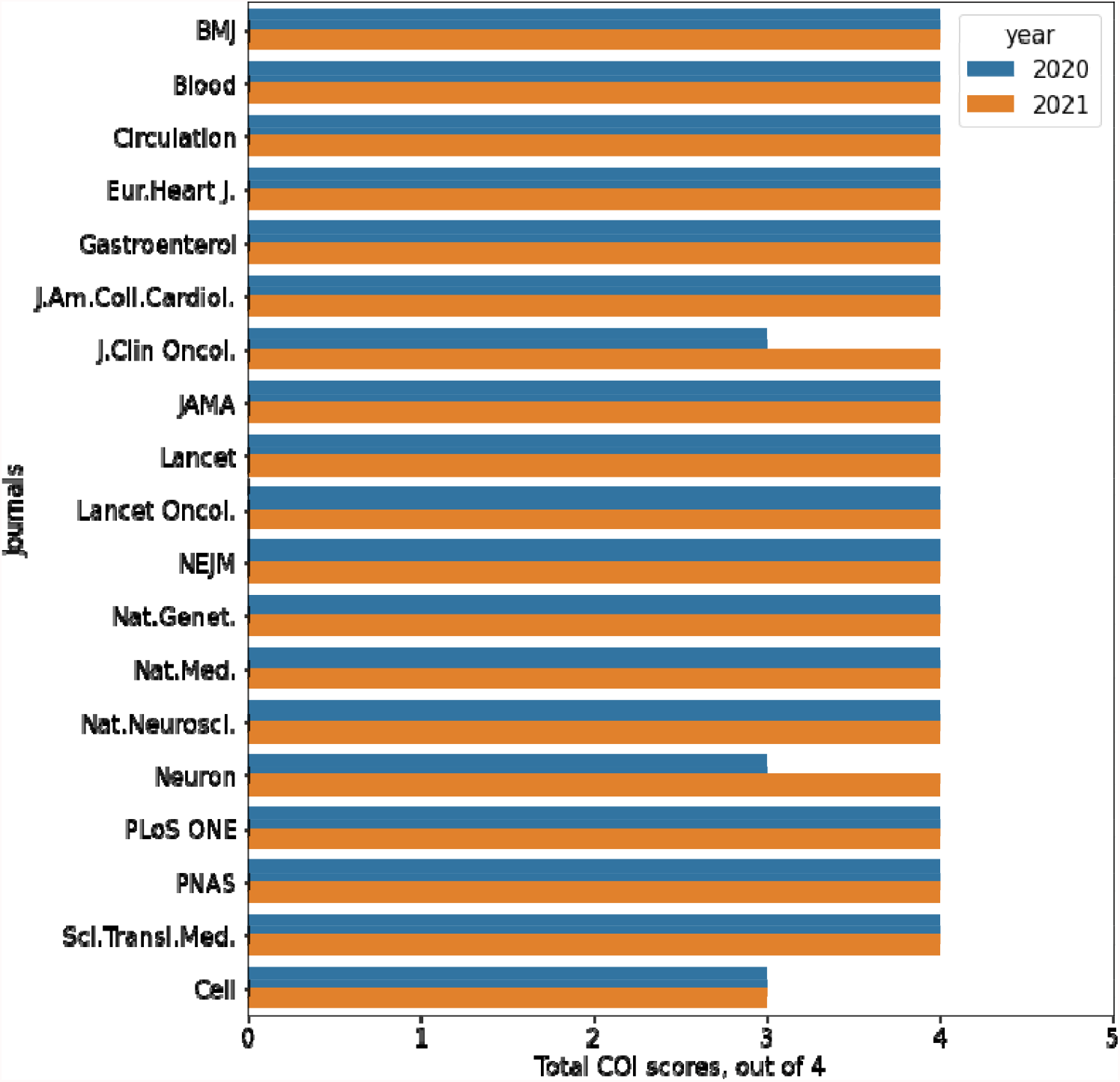
Scores of high-ranking health and medical sciences journals requirements for authors to disclose conflict of interests (COIs) using the four standards in the International Committee of Medical Journal Editors (ICMJE) disclosure form. Scores were graded twice using policies dated in February 2020 (before the COVID-19 pandemic) and in May 2021 (during the COVID-19 pandemic). Eur.Heart J: European Heart Journal; Gastroenterol: Gastroenterology; J.Clin Oncol: Journal of Clinical Oncology; J.Am.Coll.Cardiol: Journal of the American College of Cardiology; Nat.Genet: Nature Genetics, Nat.Med: Nature Medicine; Nat.Neurosci: Nature Neuroscience; PNAS: Proceedings of the National Academy of Sciences; Sci.Transl.Med: Science Translational Medicine; BMJ: The British Medical Journal; JAMA: Journal of the American Medical Association; Lancet Oncol: The Lancet Oncology; and NEJM: The New England Journal of Medicine.

## DISCUSSION

The transparency and openness standards in the policies of high-ranking health and medical sciences journals were low before and during the COVID-19 pandemic. We found limited improvement in open science standards between evaluations, with most (58%) journals making no change to their policies. The preregistration of study protocols and analysis plans and encouraging replication studies were the standards in most need of improvement. In May 2021, ten journals were signatories of the TOP guidelines, with *PNAS* signing after the first evaluation. *PLoS One* was the only journal to accept registered reports, and open science badges were not used by any journals. The majority of journals endorsed ICMJE standards for disclosing COIs.

A lack of transparency in journal policies has been found across biomedical specialties^1^. Low engagement with transparent and open science standards in journal policies have been documented in research using the TOP guidelines to assess pain journals, sports science journals, and sleep and chronobiology journals. Those studies similarly found a limited requirement for authors to preregister their studies or analytic plans, and no journals assessed encouraged replication studies^29–31^.

Incentives can improve engagement with transparency and openness standards^43^. Adopting open science badges may provide a valuable incentive for scientists to practice openly^43^. In a study by Kidwell et al., the authors found that four prominent psychology journals had less than 3% of articles reporting publicly accessible data before introducing open science badges. After these journals adopted open science badges, the same journals had nearly 40% of articles reporting publically accessible data^43^. Cashin et al.^29^ and Spitschan et al.^30^ found that no journal they assessed mentioned the use of open science badges, which was also the case for the health and medical sciences journals evaluated in our study.

In our study, a greater proportion of health and medical journals were TOP signatories by May 2021 (10/19) compared to the pain journals assessed by Cashin et al. in May 2019 (3/10). A positive correlation between being an ICMJE-member journals, using data-sharing statements, and expressing an intention to share individual patient data has been demonstrated previously demonstrated^44^. Future studies should explore whether being a TOP signatory improves the transparency and openness standards of journal policies.

TOP scores could be used to encourage journals to improve their policies. Researchers may be incentivized to submit their studies to journals that have high TOP scores, and the public may have more trust for research from such journals. However, this would require consensus for using such a metric. The journal *Epidemiology* declined to sign TOP with concerns that there was a lack of evaluation or revision over time, a ‘one-size’ application, and a disproportionate weight on the reproducibility of a finding compared with the accuracy of the research^45^. Furthermore, there is a risk of journals having variable degrees of compliance with TOP guidance in practice, as demonstrated by Goldacre et al., who found extensive breaches to CONSORT guidelines that journals endorsed in their policies^46^. These concerns could be ameliorated by COS regularly reviewing journals’ promotion and concomitant adherence to the TOP guideline and by reviewing the wording of its guideline to ensure that it may be applied to a wider range of publication types.

Journal policies that do not encourage or mandate transparent and open research reporting are likely attributable to multiple factors. To date, journals have prioritised positive results over replicability^47^. The current academic publishing ecosystem incentivises quantity over quality, and as a result, researchers may not have had formal training in the tools to practice openly. While researchers may choose to use online repositories to deposit code and data, the responsibility for sharing access to study materials will remain with authors until journal policies require data citation and sharing prior to publication. If health and medical sciences journals mandate researchers to adhere to the transparency and openness standards of the TOP guidelines, this may incentivise researchers to upskill and improve the conduct and reporting of their research. Editors should use available resources to guide recommended policy wording for improved open science practice^48^. Ongoing efforts encouraging journals to improve their transparency and openness standards will require further auditing, which would likely be facilitated if COS kept data on when journals improved their TOP scores.^49,50^

Since the COVID-19 pandemic, research stakeholders have encouraged and supported improved open science practices for research specific to the pandemic^32,33,51–54^. Despite these efforts, our study showed little improvement in the openness and transparency standards of high-ranking health and medical journals during the COVID-19 pandemic. Due to the multitude of factors that may contribute to a journal’s decision to change its policies, it is difficult to determine which improvements are a direct result of the COVID-19 pandemic.

### Strengths and limitations

While the TOP guideline is a valuable tool for assessing journals’ transparency and openness standards, there are some limitations. Firstly, the presence of imprecise and undefined terms when describing the required degree of adherence to a standard, such as ‘should’, ‘strongly encourage’, ‘recommend’ or ‘expect’, can lead to difficulties in allocating scores. To mitigate this, we conducted the scoring independently in dual and discussed scoring with a third study author to resolve differences in scoring. Our study assessed the policies of journals and not adherence of their published articles to these policies. The TOP guideline may not be applicable or flexible when examining the policies of specialized journals, such as the *Cochrane Database of Systematic Reviews*, which we excluded from our study. Nevertheless, the TOP guideline applies equally to journals that publish all study designs and provides a useful overview of how journals can improve the quality of research it publishes.

### Future research

Our methods can be replicated by others to assess the transparency and openness standards of journals in different research fields. This information can be provided to journal editors to encourage them to improve their policies. Future research should review the degree of transparency of research published in journals to assess compliance with journal policies. Furthermore, the impact of poor transparency and open science practices on individual and societal health outcomes should be studied to demonstrate its real-world consequences.

## Conclusions

We found that the 19 highly ranked health and medical sciences journals had minimal requirements for transparency and openness standards in their policies. During the COVID-19 pandemic, nominal improvements in the journal policies were observed. As the primary gatekeepers of research and evidence dissemination that impacts individual and societal health outcomes, journal policies should be regularly reviewed and improved to reflect the ongoing need for transparent and open research.

## Acknowledgements

We acknowledge all members of the Open Pain Research Appraisal and Advocacy (OPeRA) group (https://osf.io/h239s/) for providing the idea for conducting this study. We thank Assistant Professor Sean Grant for providing clarity on scoring. No funding was provided or obtained for this study.

## Author contributions

GCR prepared the study protocol with HL. DN extracted the ranking of health and medical journals from Google Scholar. ADG, EH, CJ, GT, AGC, HL, DN and ECT extracted and scored journal policies. GCR resolved disagreement of TOP scores, analysed the data, and created the code and figures. GCR, HL, and DN provided supervisory support. ADG and GCR wrote the first draft manuscript, and all authors contributed, edited, and agreed to submit the final manuscript for publication. Authorship for EH, CJ, and GT (audit 1) and AGC, HL, DN, ECT (audit 2) was determined alphabetically.

