## Supplementary material for "Open science policies of medical and health sciences journals before and during the COVID-19 pandemic: a repeat cross-sectional study": in Supplement 1

Summary of scores for the eight TOP standards

**Data citation**

Journal policies had a median score of 0 (IQR: 0-2) in February 2020 for data citation, which increased to a median of 1 (IQR: 0-2) in May 2021. Most (89%; n=17) journals did not change their score for data citation. *Gastroenterology* and the *European Heart Journal* improved their scores from 0/3 in February 2020 to 2/3 in May 2021. In both evaluations, three journals scored one out of three for data citation (Table 1), meaning that their policy “described citation of data in guidelines to authors with clear rules and examples.” In May 2021, eight journal policies scored two out of three, stating that they “required appropriate citation for data and materials”. Eight journals scored zero out of three, meaning they did not mention any requirements for citing data. No journal policy scored three out of three in either timepoint for data citation, meaning that the journals did not verify citation for data and materials before publication.

**Data transparency**

At both time points, the 19 journal policies had a median score of 1 (IQR: 0.5-2 in February 2020 policies; IQR: 1-2 in May 2021) for data transparency, meaning that authors “must state whether or not data are available, requiring a data availability statement satisfies this level”. The increased lower IQR was driven by the *European Heart Journal*, *The Lancet*, and *The Lancet Oncology*, who each scored 0/3 in February 2020 and 1/3 in May 2021. Most (84%; n=16) journals did not change their scores for data transparency. In May 2021, two journal policies scored zero out of three and eight scored one out of three for data transparency. In both evaluations, nine journal policies scored two out of three for data transparency, meaning that their policy states that authors “must have publically available data, or an explanation why ethical or legal constraints prevent it”. No journal policy scored three out of three points for data transparency, meaning that the journals did not require articles to “have publically available data and must be used to computationally reproduce or confirm results before publication”.

**Code transparency**

Scores for code transparency increased from a median of 0 (IQR: 0-2) in February 2020 to a median of 1 (IQR: 0-2) in May 2021. The *European Heart Journal*, *Gastroenterology*, and *PLoS ONE* improved their coding policies scoring zero before the COVID-19 pandemic to scoring one in May 2021. *Cell* and *Neuron* changed the wording of their policy from “Code must also be provided to editors and peer reviewers at the time of submission for the purposes of evaluating the manuscript” scoring 3/3 in February 2020, to requiring authors to deposit their code before publication, but not reproducing the code before publication, scoring 2/3 in May 2021. Overall, most (74%; n=14) journals did not change their policies on code transparency. In May 2021, seven journal policies scored zero and four scored one for code transparency, meaning that their policy required authors to “state whether or not code is available”. In May 2021, six journal policies scored two out of three for code transparency, meaning that authors “must have publically available code, or an explanation why ethical or legal constraints prevent it”. In May 2021, two journal policies scored three out of three for code transparency, meaning that articles “must have publically available code and must be used to computationally reproduce or confirm results prior to publication”.

**Materials transparency**

Journal policies scored a median of 1 in both February 2020 (IQR: 0-2) and May 2021 (IQR: 1-2) for materials transparency, meaning that authors “must state whether or not materials are available, a materials availability statement satisfies this level”. Most (84%; n=16) journals did not change their policies on materials, but the *European Heart Journal*, *The Lancet*, and *The Lancet Oncology* improved their score from 0/1 in February 2020 to 1/1 in May 2021. In May 2021, four policies scored zero and seven scored one out of three for materials transparency. In May 2021, eight journal policies scored two out of three, meaning that articles “must have publically available materials or an explanation why ethical or legal constraints prevent it”. No journal policy scored three out of three, meaning that articles did not need to make materials publically available and “computationally reproduce or confirm results prior to publication”.

**Design and analysis transparency**

In both evaluations, journal policies had a median score of 1 for design and analysis transparency (IQR: 0.5-2 for 2020; 1-2 for 2021). *Gastroenterology* improved its score from 0/3 in 2020 to 1/3 in 2021. Most (95%; n=18) journals had no difference in their design and analysis transparency standard scores. In May 2021, four policies scored zero and did not mention reporting guidelines; six journals scored one out of three points for design and analysis transparency, meaning that the policy “articulates design transparency standards”; nine journals scored two out of three, meaning that the policy “requires adherence to design and transparency standards for review and publication”. No journal policy required and enforced “adherence to design transparency standards for review and publication”.

**Preregistration of studies**

Journal requirements for study preregistration improved between the two evaluations, with policies scoring a median of 0 (IQR: 0-1) in February 2020 and a median of 1 (IQR: 0-1) in May 2021. One journal, *Gastroenterology,* strengthened their policy for study preregistration from 0/3 in February 2020 to 1/3 in May 2021. Most (95%, n=18) journal policies had no changes in the requirements for study protocol preregistrations. In May 2021, nine journal policies scored zero and ten scored one out of three points, meaning that the journal required articles to “state if work was preregistered”. No journal policy scored two or three for study preregistration, meaning that the policies did not require and verify adherence to the preregistered plan.

**Preregistration of analysis plans**

Similar to study preregistration, journal policies for the preregistration of analysis plans increased from a median of 0 (IQR: 0-1) in February 2020 to 1 (IQR: 0-1) in May 2021. *Gastroenterology* improved its policy, scoring 0/3 in February 2020 to 1/3 in May 2021. Most (95%, n=18) journal policies had no changes in the requirements for preregistration of analysis plans, with nine policies scoring zero and ten scoring one out of three in May 2021, meaning that “articles will state if work was preregistered with an analysis plan”. No journal policy scored two or three for the preregistration of analysis plans.

**Replication**

The policies of journals regarding the submission of replication studies improved from a median score of 0 (IQR: 0-1) in February 2020 to a median score of 1 (IQR: 0-1) in May 2021. *Gastroenterology* and *PLoS ONE* improved their policies, scoring 0/3 before the COVID-19 pandemic to 1/3 in May 2021. Most (89%; n=17) journals did not change their policies for submitting replication studies. In May 2021, nine policies scored zero, and ten policies scored one out of three, meaning that they “encouraged the submission of replication studies”. No journal policy scored two or three for replication studies, meaning that the journals did not “review replication studies blinded to results” or use “registered reports for replications as a regular submission option”, respectively.
